# A culturally and personally tailored cooking application improves glycemic control in type 2 diabetes

**DOI:** 10.64898/2025.12.14.25342210

**Authors:** Shoko Takei, Kazuki Hamada, Naru Shimizu, Momoe Sakurai, Tetsuya Nojiri, Naoya Yahagi

## Abstract

**Aims:** To evaluate the effectiveness of the Oishi-kenko mobile cooking application in improving glycemic control, body weight, and dietary behaviors among individuals with type 2 diabetes without professional intervention.

**Methods:** This 12-week observational study was conducted entirely online. Participants were recruited via the Oishi-kenko website, internet advertisements, and Tsukuba City public relations channels. Of 24,671 website visitors, 214 installed the app, and 65 were included in the final analysis; HbA1c data were available for 24 participants. The app provided personalized, culturally tailored recipe suggestions based on user profiles and dietary standards. Body weight and HbA1c were assessed at baseline and every 4 weeks, and dietary habits were evaluated using the Brief-type Self-administered Diet History Questionnaire (BDHQ).

**Results:** Mean HbA1c decreased from 6.40% at baseline to 6.12% after 12 weeks (-0.28%, P<0.05). Among participants with baseline HbA1c >7%, the reduction was -0.63%. BMI declined from 23.47 to 23.28 overall, with greater reduction among those with baseline BMI ≥25. More frequent app-based cooking and higher baseline HbA1c predicted greater improvement. BDHQ analyses showed reduced intake of salty condiments, noodle soup, and fatty meats, alongside healthier eating behaviors. Over 80% of participants reported improved dietary habits.

**Conclusions:** Use of the Oishi-kenko app was associated with improved glycemic control, modest weight loss, and healthier eating patterns in individuals with type 2 diabetes. These findings support the potential of culturally tailored, stand-alone dietary support applications as scalable tools for diabetes self-management.

(The trial registry number: UMIN-CTR, ID: R000053861)

**Highlights:**

- Oishi-kenko app use significantly reduced HbA1c and BMI in individuals with type 2 diabetes.
- The study was conducted fully online, from recruitment to data collection, without human intervention.
- Participants adopted healthier dietary behaviors, including reduced intake of salty condiments, noodle soup, and fatty meats, along with more mindful eating practices.
- The app shows promise as a stand-alone, culturally tailored digital tool for diabetes self-management without professional intervention.

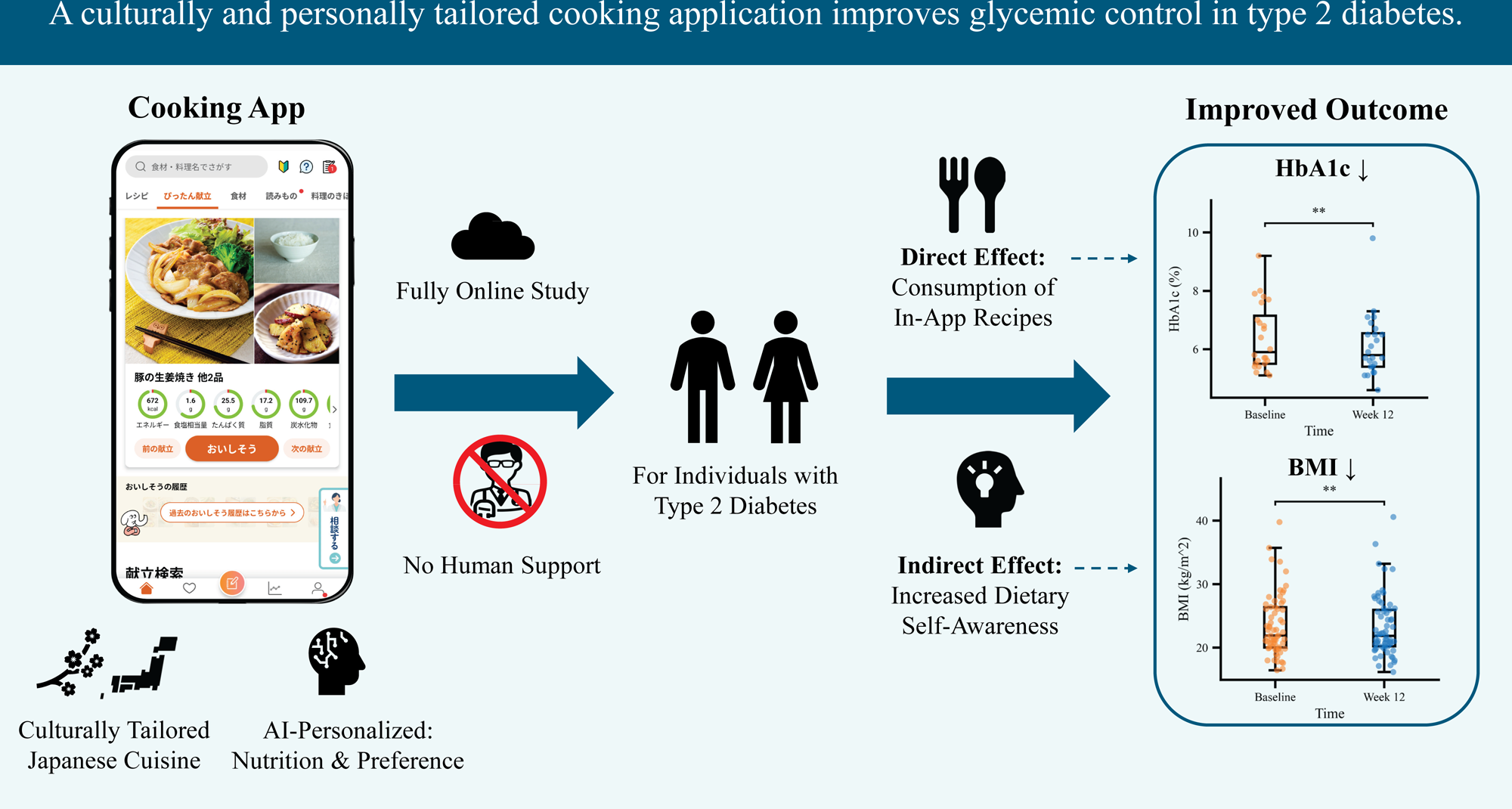

## Introduction

Diabetes mellitus is a prevalent metabolic disorder characterized by chronic hyperglycemia resulting from impaired insulin secretion, insulin action, or both ^1^. In 2021, an estimated 529 million people worldwide were living with diabetes, and this number is projected to exceed 1.3 billion by 2050 ^2^. Poor glycemic control is associated with the development of microvascular and macrovascular complications, including retinopathy, nephropathy, neuropathy, and cardiovascular disease, all of which substantially increase morbidity and mortality ^3^. Accordingly, optimizing glycemic management remains a central goal in the treatment of type 2 diabetes.

Standard therapeutic strategies for type 2 diabetes involve a comprehensive approach that combines dietary modification, regular physical activity, and pharmacological therapy. For example, the American Diabetes Association (ADA) recommends diabetes self-management (DSM) practices such as maintaining a healthy diet, engaging in physical activity, avoiding tobacco use and excessive alcohol consumption, adhering to prescribed medications, and monitoring blood glucose levels ^4^. Lifestyle interventions are particularly critical, as dietary intake and physical activity directly influence both insulin sensitivity and glycemic variability. However, despite their importance, adherence to lifestyle recommendations remains challenging for many patients, highlighting the need for accessible and sustainable support systems to facilitate behavioral change.

In recent years, digital health interventions — particularly smartphone-based applications — have emerged as promising tools to support diabetes self-management and reinforce lifestyle modification. Such applications can deliver real-time feedback, educational resources, and interactive support, thereby enhancing patient engagement and adherence ^5–8^. However, many previous studies have included human components, such as in-person counseling or coaching, which may limit scalability and generalizability.

The Oishi-kenko application is a smartphone-based platform designed to support users in the efficient preparation of nutritious and appetizing Japanese home-cooked meals ^9^. By providing recipe suggestions tailored to individual preferences and nutritional needs, the app aims to promote healthier eating habits without compromising taste or convenience. Its culturally adapted design offers a practical approach to supporting dietary modification among Japanese individuals with type 2 diabetes.

In this study, we evaluated the effect of the Oishi-kenko app on glycemic control in individuals with type 2 diabetes, focusing specifically on outcomes achieved without human intervention.

## Methods

### Study aim

This was an observational study to examine whether patients with type 2 diabetes, and individuals interested in the Oishi-kenko app’s dietary topics related to type 2 diabetes, could improve glycated hemoglobin (HbA1c) levels and body weight through app use.

### Study design

This study was conducted entirely online, from participant recruitment to research content explanation and data collection. Participants used the Oishi-kenko app freely for 12 weeks and completed questionnaires on topics such as dietary intake using the BDHQ (brief-type self-administered diet history questionnaire) ^10^ and smartphone use at the beginning and end of the study. Body weight and HbA1c were collected every four weeks from the start of this study. This study was approved by the Ethics Committee of the University of Tsukuba Hospital (Approval No.: R03-266) and was registered with the UMIN Clinical Trials Registry (UMIN-CTR, ID: R000053861).

### Data collection

Participants were recruited using the Oishi-kenko website and internet advertisements, and with the cooperation of Tsukuba City, the city’s public relations magazine and website. During the recruitment period from May 2022 to September 2023, 214 people agreed to participate in this study and downloaded app. 171 people responded to the first questionnaire. 93 people responded to the 12th week questionnaire at the end of this study. Participants who responded to the final questionnaire but did not have verifiable data registered, and participants who did not access the app after the initial download were excluded. Finally, 65 participants were analyzed. However, HbA1c was analyzed in 24 people who had entered usable data.

### App features

The Oishi-kenko application is a smartphone-based platform designed to support users in the efficient preparation of nutritious and appetizing Japanese home-cooked meals ^9^. It has functions for searching for meal recipes, suggesting daily meal plan which combine recipes, and recording life logs such as meals, weight, and blood pressure. The app collects information such as age, sex, recipe viewing history, and favorite registrations, which it uses to predict user preferences. Based on these data, artificial intelligence (AI) suggests recipes and menus that meet nutritional standards calculated from the user’s profile. These recipes are suggested based on Japan’s Dietary Reference Intakes for 2020 (Dietary Reference Intakes for Japanese, 2020 version). These recipes and menus were personalized, considering the individual’s height, weight and comorbidities ^9^.

### Statistical analyses

Comparisons between baseline and 12-week data were performed using the two-tailed Wilcoxon signed-rank test, as the assumptions of normality and homogeneity of variance required for the t-test were not satisfied. The corresponding p-values were calculated. Statistical significance was defined as *P < 0.05, with trends indicated by †P < 0.1 and stronger significance by **P < 0.01. All statistical analyses were conducted using the SciPy library (version 1.13.1) in Python (version 3.11).

## Results

### Participant flow and retention

**Figure 1A** illustrates the flow of participants. A total of 24,671 people visited the recruitment website, of which 654 consented to participate in this study. 394 were able to verify their identity, and 214 installed the app. 171 participants responded to the first questionnaire, 113 to the 4^th^ week questionnaire, 101 to the 8^th^ week questionnaire, and 93 to the end-of-study 12^th^ week questionnaire. Of these, 65 participants could be analyzed.

**Figure 1.**
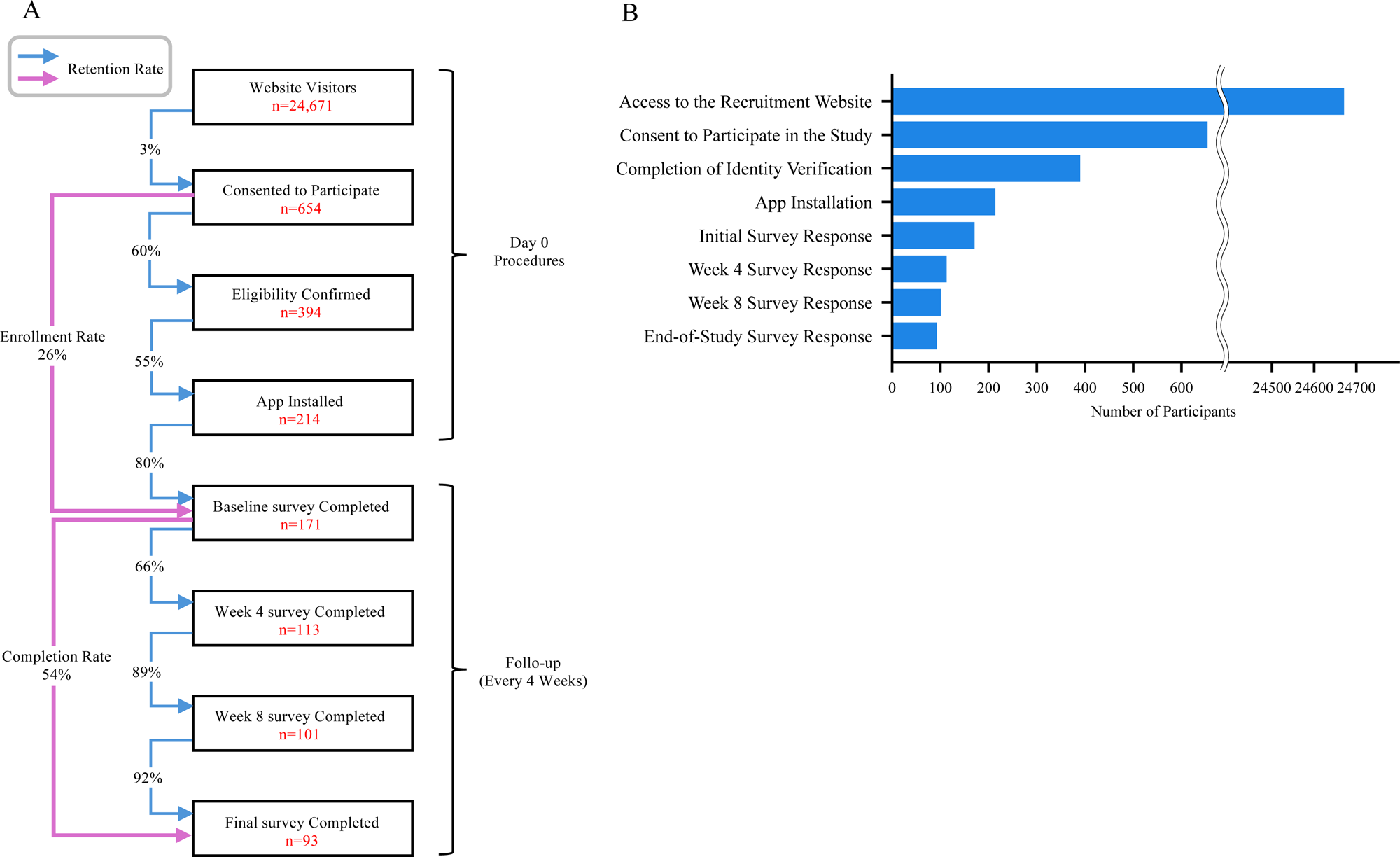
Participant flow and retention rates at each stage. (A) Flowchart illustrating the participant flow from 24,671 website visitors to consent for study participation, identity verification, app installation, and completion of each survey. The number of participants at each stage and the **retention** rate to the next stage are shown. (B) Bar chart showing the number of participants who completed each procedure from website visit to final survey response.

After obtaining consent for this study, only 60% of participants were able to confirm their identity. Furthermore, only 55% of participants installed the app after confirming their identity (**Figure 1B**). After completing the first questionnaire, only 66% of respondents completed the 4th week questionnaire. The retention rates for app installation, identity verification, and responses to the 4th week questionnaire were low.

### Baseline characteristics and app use

**Table 1** shows the baseline characteristics. Of the 65 participants, 16 were male and 49 were female, with a mean age of 44.51 ± 3.02 years. The mean weight was 61.15 ± 3.56 kg, BMI was 23.47 ± 1.17, and HbA1c was 6.4 ± 0.28% at the start of this study.

**Table 1.**
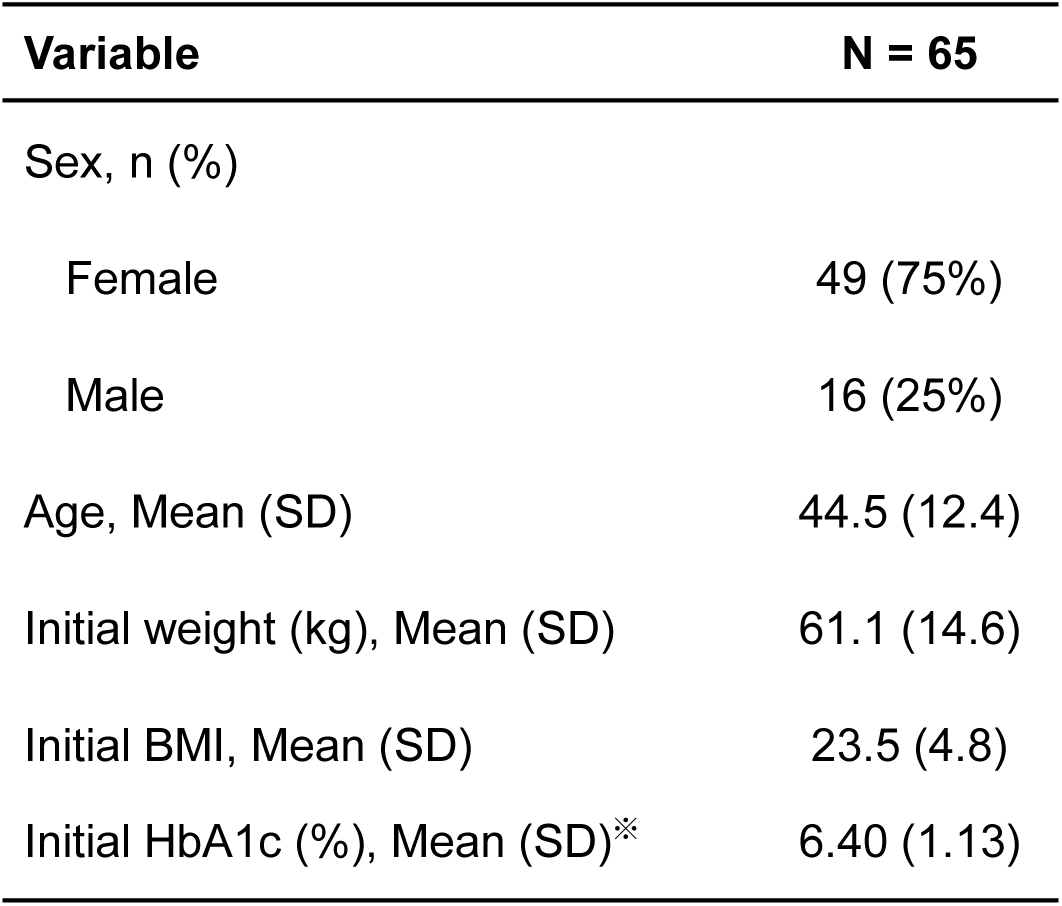
Baseline characteristics of participants. Of the 65 participants, 16 were male and 49 were female, with a mean age of 44.51 ± 3.02 years. HbA1c values were calculated based on N = 24.

The average number of days per week that participants used the app was 1.64, and 34 participants (52%) used the app at least once a week. There were no restrictions on the frequency of app use, so usage varied among participants (**Figure 4**).

### Changes in HbA1c and BMI after 12 weeks

**Figure 2** shows the changes in HbA1c levels and BMI after 12 weeks. The mean HbA1c at baseline was 6.4%, which significantly decreased to 6.12% after 12 weeks (**Figure 2A**). The overall reduction was 0.28%, but among participants with baseline HbA1c ≥7% (a group not meeting the Japan Diabetes Society’s target of <7% for complication prevention), the reduction was 0.63% (**Figure 2B**), indicating that greater improvements were seen in those with higher baseline HbA1c.

**Figure 2.**
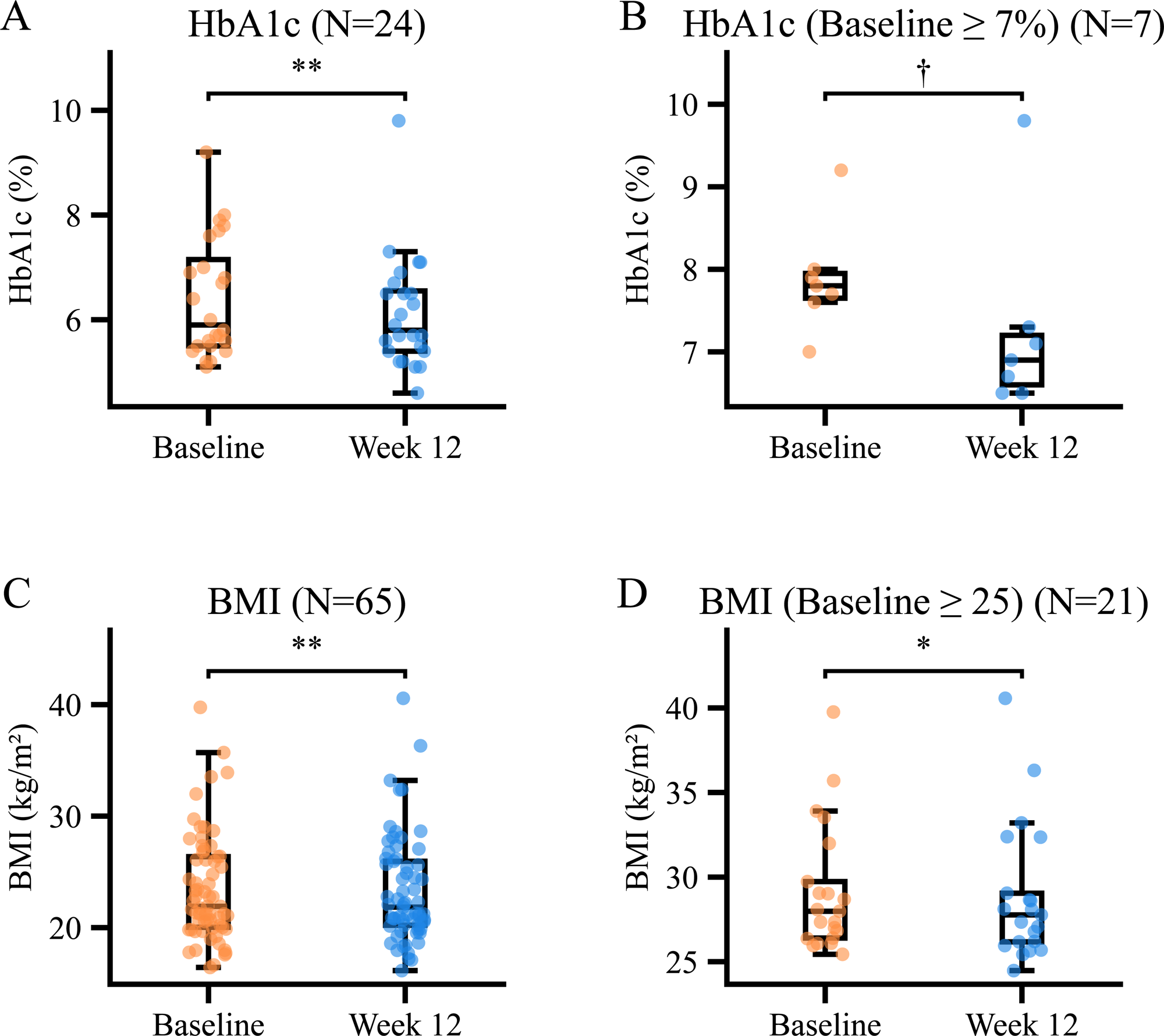
Changes in HbA1c and BMI after 12 weeks. (A) Individual plots and boxplots of HbA1c at baseline and 12 weeks for participants with HbA1c data available at both time points (N=24). (B) Individual plots and boxplots of HbA1c at baseline and 12 weeks for participants with a baseline HbA1c ≥7% (N=7). (C) Individual plots and boxplots of BMI at baseline and 12 weeks for all participants (N=65). (D) Individual plots and boxplots of BMI at baseline and 12 weeks for participants with a baseline BMI ≥25 (N=21). Comparisons between baseline and 12-week data were performed using the two-tailed Wilcoxon signed-rank test, as the assumptions of normality and homogeneity of variance required for the t-test were not satisfied. Statistical significance was defined as *P* < 0.05, with trends indicated by †*P* < 0.1 and stronger significance by **P* < 0.01.

BMI also decreased significantly, from 23.47 at baseline to 23.28 at 12 weeks (**Figure 2C**). For those with a baseline BMI ≥25, which is classified as obese by the Japan Society for the Study of Obesity, BMI decreased from 29.16 to 28.85, again showing greater improvement in participants with higher baseline BMI (**Figure 2D**).

Multiple regression analysis, adjusted for potential confounding variables, revealed that the frequency of cooking using the app and baseline HbA1c levels significantly affected changes in HbA1c, thereby evaluating the relationship between app usage and HbA1c improvement (**Table 2**).

**Table 2.** Multivariate analysis of the association between app-based cooking frequency and change in HbA1c. Adjusted R² = 35.2%. It revealed that the frequency of cooking using the app and baseline HbA1c levels significantly affected changes in HbA1c, thereby evaluating the relationship between app usage and HbA1c improvement.

| Variable | Coefficient | Standard Error | p value |
| --- | --- | --- | --- |
| Intercept | 1.914 | 0.658 | .009 |
| App-Based Cooking Frequency<br>(days per week) | -0.0896 | 0.042 | .046 |
| Sex | -0.258 | 0.196 | .205 |
| Age | -0.0009 | 0.009 | .919 |
| Initial HbA1c | -0.195 | 0.090 | .043 |
| Baseline Cooking Frequency<br>(days per week) | -0.087 | 0.037 | .030 |

### Changes in dietary habits after 12 weeks

To explore mechanisms underlying the app’s effects, BDHQ analyses were performed at baseline and after 12 weeks. The number of participants who reported drinking noodle soup frequently decreased (**Figure 3A**), while those who reported that their home-cooked meals were slightly less salty increased (**Figure 3B**).

**Figure 3.**
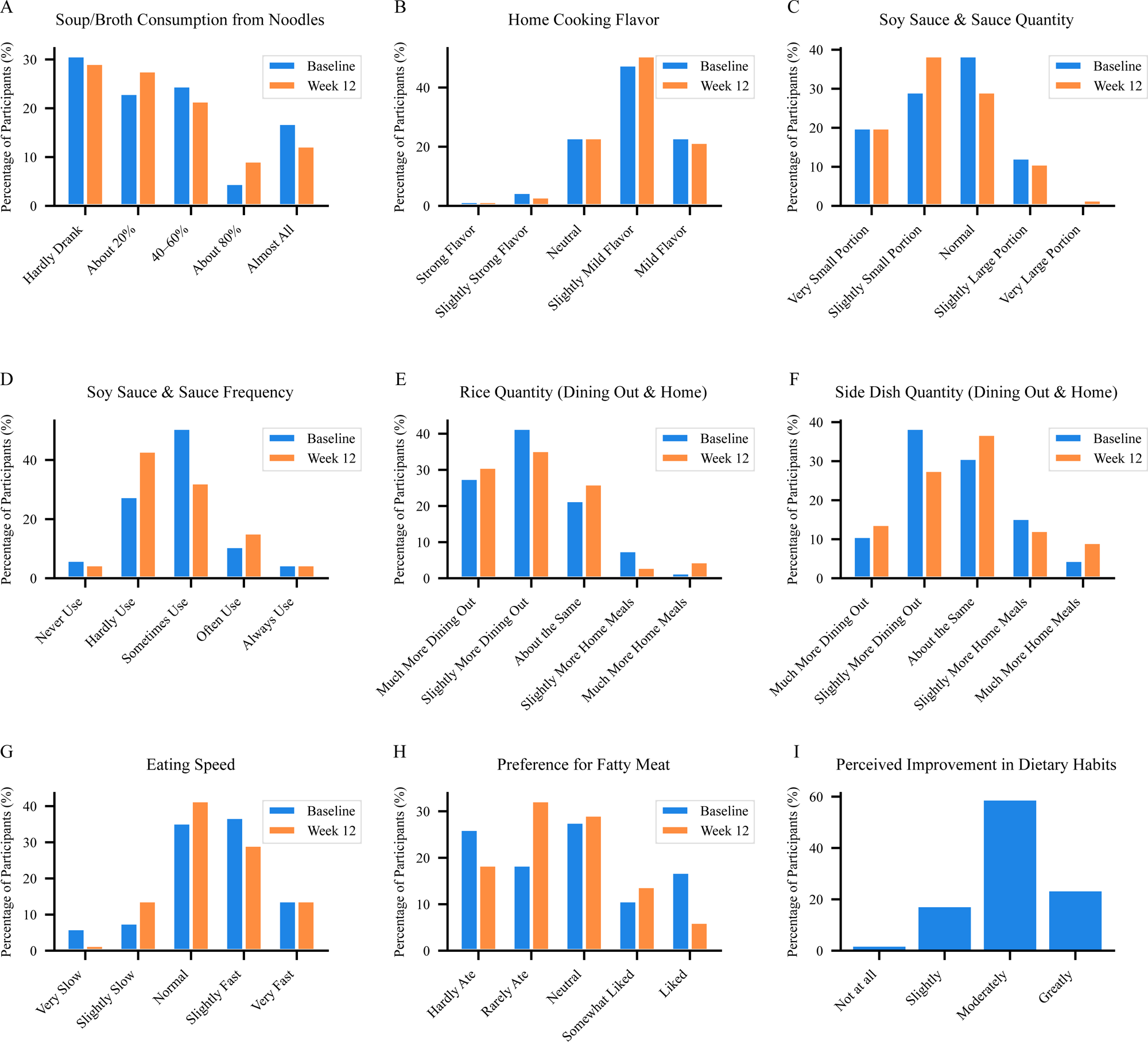
Changes in dietary habits after 12 weeks. (**A**-**H**) Changes in participants’ dietary habits based on the brief-type self-administered diet history questionnaire (BDHQ) administered at baseline and 12 weeks. Each graph shows the percentage of respondents at baseline (blue) and 12 weeks (orange). (**I**) Results of self-assessment on the improvement of dietary habits after 12 weeks of app use, based on an original questionnaire.

**Figure 4.**
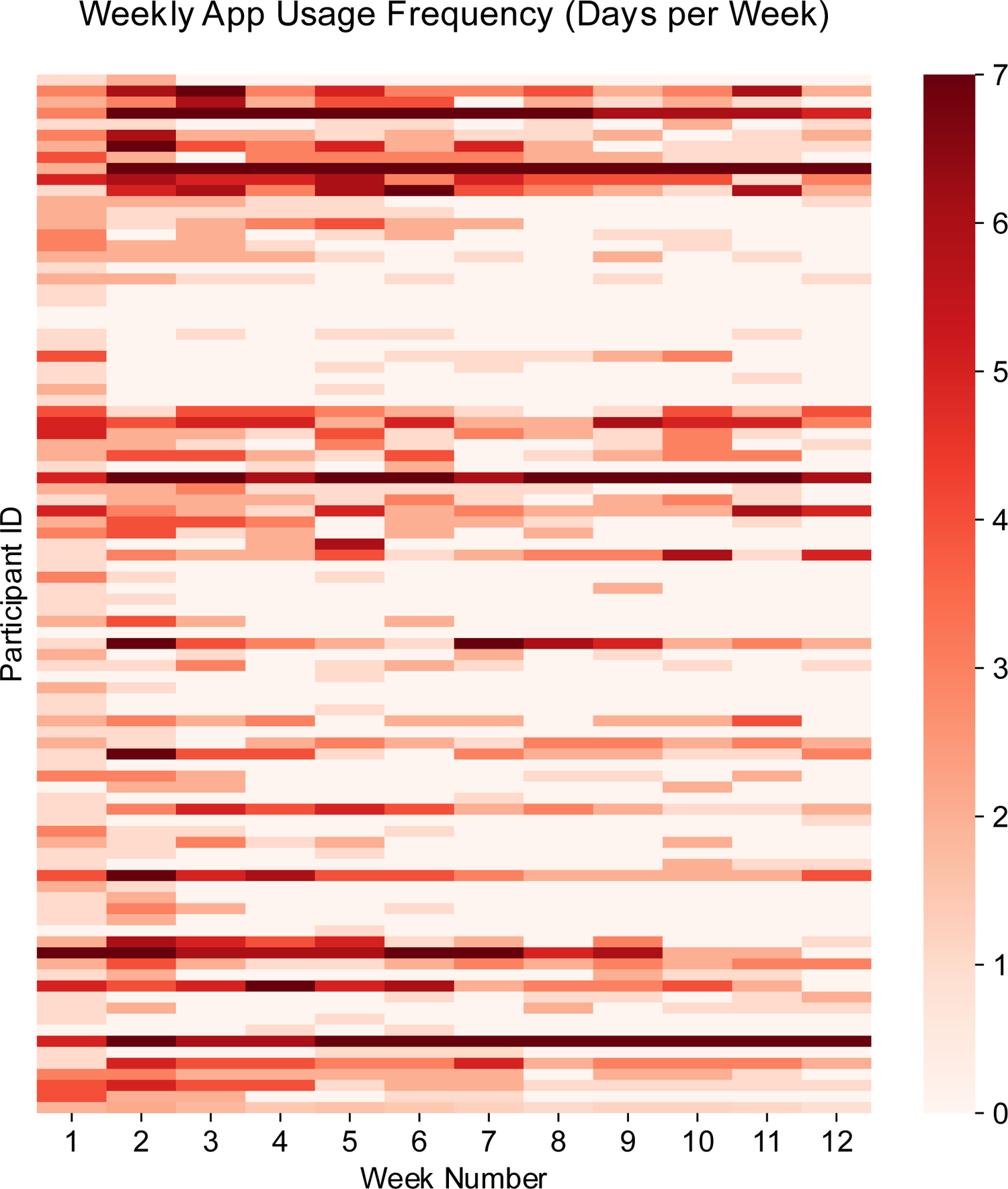
Weekly app usage frequency. The average number of days per week that participants used the app was 1.64, and 34 participants (52%) used the app at least once a week.

Similarly, the percentage of participants who reported using less soy sauce or sauce increased (**Figure 3C**), and those who reported very rare use also increased (**Figure 3D**). The proportion of participants who ate slightly more rice when eating out decreased (**Figure 3E**), and the proportion who reported larger side dish portions when eating out also decreased (**Figure 3F**).

Eating behaviors also improved: more participants reported eating at a slightly slower or normal speed (**Figure 3G**), and the proportion who reported eating little fatty meat increased (**Figure 3H**). Furthermore, over 80% of participants answered that using the app had somewhat or very much improved their eating habits (**Figure 3I**).

## Discussion

In this study, use of the Oishi-kenko mobile application was associated with significant improvements in glycemic control and body weight among individuals with type 2 diabetes. HbA1c decreased by 0.28% overall and by 0.63% in participants with higher baseline HbA1c (>7%), and BMI also declined modestly over 12 weeks. These results suggest that the app can support lifestyle modification and improve clinical outcomes without direct human intervention.

The mechanisms underlying these benefits appear to involve both direct and indirect pathways. On the one hand, participants who used the app more frequently, particularly for cooking, experienced greater reductions in HbA1c, indicating that repeated engagement with tailored recipe suggestions and meal planning directly facilitated healthier dietary choices. On the other hand, the app also influenced participants’ awareness of their eating patterns. Analyses of the BDHQ revealed decreased consumption of salty condiments, noodle soup, and fatty meats, alongside increased reports of mindful eating behaviors such as slower eating speed and portion control. More than 80% of participants reported subjective improvement in eating habits, suggesting that the app functioned not only as a recipe provider but also as a behavioral catalyst that enhanced dietary self-awareness.

To our knowledge, this is the first study in which recruitment, questionnaire completion, and app use were all conducted entirely online. Conducting the trial fully remotely allowed us to examine the effectiveness of dietary management using the app in a real-world setting. A meta-analysis of 10 studies evaluating apps for type 2 diabetes reported a mean HbA1c reduction of 0.55% among app users; however, 8 of these 10 studies included additional feedback by health care professionals ^11^. By contrast, our study demonstrated improvements with an app-only intervention, suggesting that dietary support applications can provide benefits without direct professional involvement.

Our findings also compare interestingly with studies of other digital interventions. For example, BlueStar, a smartphone app designed to improve diabetes self-management, significantly reduced HbA1c levels in an early trial ^12^ and became the first mobile prescription therapy approved by the U.S. Food and Drug Administration. However, a subsequent multicenter randomized controlled trial reported no significant difference in glycemic control between the intervention and control groups, underscoring the variability of outcomes across settings ^13^. Similarly, the DIABEO application, which integrates insulin dose calculation with real-time monitoring, achieved significant HbA1c reduction in a randomized trial, but this effect was strongly dependent on daily app use ^14^. These findings highlight that the intensity of engagement remains a key determinant of effectiveness across digital health tools.

The global penetration of smartphone technology further emphasizes the potential impact of such interventions. As of April 2025, 7.21 billion people — equivalent to 89% of the world’s population — own a smartphone. This widespread availability makes digital health solutions more convenient and accessible than face-to-face medical care in many contexts. At the same time, sustained engagement remains a challenge: more than two-thirds of individuals who download mobile health applications use them only once ^15^. Identifying strategies to promote continued use and integrate digital tools into daily life will therefore be essential for maximizing their long-term benefits. In this regard, the Oishi-kenko app incorporates several features that may encourage ongoing use, including personalized menu suggestions tailored to user preferences and nutritional needs, as well as an emphasis on taste and convenience alongside health. By providing practical, culturally familiar recipes, it distinguishes itself from applications that focus primarily on calorie tracking or education, which may explain the high levels of user-reported improvement in eating habits. Together, these elements lower psychological barriers to dietary change and help sustain user interest over time, thereby enhancing long-term engagement and effectiveness compared with generic diet-tracking applications.

Several limitations of our study should also be acknowledged. The sample size was modest, and HbA1c data were available for only a subset of participants, which limited statistical power. Retention was also relatively low, with fewer than half of initial registrants completing the study, raising the possibility of selection bias. Moreover, the average frequency of app use was only 1.6 days per week, suggesting that stronger effects might be observed with higher engagement. In addition, because this was an observational study without a control group, causal inference is not possible. Finally, as the study was conducted in Japan using a culturally tailored application, the findings may not be generalizable to other populations.

Despite these limitations, it is noteworthy that even with limited average usage, participants reported meaningful changes in dietary behavior and experienced modest but significant improvements in metabolic outcomes. This suggests that even small, repeated exposures to dietary prompts and personalized recipes can accumulate to produce health benefits. The scalability of app-only interventions also makes them particularly attractive in public health contexts where resources for individual counseling may be limited.

In conclusion, use of the Oishi-kenko mobile application was associated with improved glycemic control, modest weight reduction, and healthier dietary behaviors in individuals with type 2 diabetes. These effects appear to derive both from direct engagement with the app and from increased dietary self-awareness fostered by its use. Although further research with larger, controlled trials is warranted, these findings support the potential of culturally tailored, stand-alone dietary support applications as accessible tools for diabetes self-management in the digital health era.

## Data Availability

All data produced in the present work are contained in the manuscript

## Acknowledgements

This research was supported by AMED under Grant Number JP23gm1710008 (AMED-CREST) and JP23rea522010 (Healthcare Social Implementation Infrastructure Development Project) (to N. Yahagi). This research was also supported by AMED under Grant Number JP24le0110033 (to Oishi-kenko).

## Author contributions

N.Y. and T.N. conceived the study. K.H. and N.S. performed the study under the guidance of T.N. and N.Y. and analyzed the data together with S.T. T.N. and N.Y. S.T. and N.Y. co-wrote the original manuscript. All authors discussed the results and commented on the manuscript.

## Declaration of interests

The authors declare no competing financial and non-financial interests.

## Abbreviations

BMI: body mass index
BDHQ: brief-type self-administered diet history questionnaire.

## Notes

### Competing Interest Statement

The authors have declared no competing interest.

### Clinical Trial

UMIN-CTR, ID: R000053861

### Funding Statement

This study was funded by by AMED under Grant Number JP23gm1710008 (AMED-CREST) and JP23rea522010 (Healthcare Social Implementation Infrastructure Development Project) (to N. Yahagi). This research was also supported by AMED under Grant Number JP24le0110033 (to Oishi-kenko).

### Author Declarations

The Ethics Committee of the University of Tsukuba Hospital gave ethical approval for this work.

